# Prebiotic soda lowers postprandial glucose compared to traditional soda pop: a randomized controlled trial

**DOI:** 10.1101/2025.04.18.25326077

**Authors:** Colleen F McKenna, Noah Voreades, Michael J Weiser, Courtney McCormick, Ana M Valdes, Traci Blonquist, Valerie Kaden, Eunice Mah, Chad Cook

## Abstract

Inadequate dietary fiber intake and excess added sugar intake are dietary factors attributed to the rise in obesity, type 2 diabetes, and associated cardiometabolic diseases. Until recently, consumers had limited options for finding similar tasting, yet highly approachable, solutions to meet intake recommendations for added sugars and dietary fiber. Modern sodas with lower sugar and supplemented with prebiotic fiber may serve as functionally beneficial alternatives to traditional sugar-sweetened beverages. The primary objective was to compare the acute effects of a prebiotic soda, (PREB: 3g sugar, 6g dietary fiber) versus traditional soda (SODA: 39g sugar, no dietary fiber), with or without a meal, on postprandial glucose in generally healthy adults. Thirty middle-aged men and women [19 F, 11 M; (mean ± standard deviation) 46.5 ± 10.4 y; 29.5 ± 2.6 kg/m^2^] consumed their assigned study product during the traditional lunch time in a free-living setting on 4-consecutive test days in a crossover design, counterbalanced by test sequence. Continuous glucose monitoring for blood glucose dynamics, visual analog scales for perceived hunger and alertness, and dietary logs for second meal behavior were all measured throughout the intervention. Glucose incremental area under the curve (iAUC) [median difference (95% confidence interval); –837 mg/min/dL (–1250, 188), p=0.032; –1690 mg/min/dL (–2790, –909), p<0.001; with and without meal, respectively] and maximum glucose concentration (C_max_) [-9 mg/dL (–27, 0), p=0.018; –36 mg/dL (–50, –22), p<0.001; with and without meal, respectively] were lower with PREB compared to SODA. PREB did not affect second meal timing nor energy intake compared to SODA. Perceived hunger or alertness were not altered by beverage type. In conclusion, a prebiotic soda is a favorable alternative to traditional soda formulations for managing postprandial blood glucose levels and maximal glucose excursion in generally healthy adults with overweight or obesity.

## INTRODUCTION

Overweight and obesity affect a substantial portion of the U.S. adult population, leading to its recognition as a critical public health issue. In 2021, an estimated 172 million adults were living with overweight or obesity, with forecasts indicating prevalence to reach 213 million adults by 2050 (1). Obesity is a known risk factor for several cardiometabolic abnormalities, including hyperinsulinemia, insulin resistance, prediabetes, type 2 diabetes, hypertension, and dyslipidemia (2, 3). Conversely, glucometabolic abnormalities, such as insulin resistance, have been shown to further advance obesity, often through mechanisms related to excess fat storage mediated by glucose and lipid metabolism, appetite regulation, and energy expenditure (4).

It is established that the causes of obesity are complex, influenced by multiple factors including genetics, lifestyle (eating habits, physical activity), socioeconomic, and environmental (5). Despite these complexities, researchers are aligned on the significant role lifestyle factors, such as dietary patterns and eating habits, play in the etiology of obesity (5). Unhealthy dietary patterns, characterized by high intakes of refined, high-glycemic load carbohydrates and low intakes of dietary fiber, play a crucial role in the development of obesity (4).

To promote health and prevent the burden of obesity and cardiometabolic risk factors, current national dietary guidelines encourage dietary patterns consisting of nutrient-dense foods and beverages across all food groups, in recommended amounts, and within calorie limits (6). While the primary focus of current dietary recommendations centers around overall dietary patterns, dietary fiber and added sugars are identified as nutrients of concern due to underconsumption and overconsumption, respectively (6).

A healthy dietary pattern limits added sugars to less than 10% of the total daily caloric intake (6). Despite that recommendation, more than half of the U.S. adult population falls short in meeting it (7). Strategies have been proposed to reduce intakes of added sugars, including reducing major sources of added sugar by means of consuming less, reducing portion sizes, and swapping food choices for lower added sugar alternatives (6).

A leading source of added sugars in the U.S. diet are sugar-sweetened beverages (SSBs), with about one-half of U.S. adults consuming at least one SSB per day during 2011 – 2014, representing about 6.5% of total daily caloric intake from SSB (6, 8). In 2020, nearly 300,000 new type 2 diabetes cases and nearly 67,000 new cardiovascular disease cases in the U.S. were attributable to SSBs consumption, representing 15.7% and 5.4%, respectively, of all incident cases (9–11).

It is clear that reducing intake of added sugars in the diet, including those provided through SSBs, is an important step to improving overall diet quality and cardiometabolic risk factors (12, 13). Indeed, diet or other zero sugar sodas that may contribute to decreases in total daily added sugar intake have been on the market for nearly half a century. However, these products dually lack the addition of prebiotic fiber—which improves cardiometabolic risk factors (14, 15), and include non-nutritive synthetic sweeteners—which are contraindicated for weight management and chronic disease risk (16). In recent years, the beverage industry has responded with an increase of ‘better for you’ beverage options, lending the way to the establishment of the “modern soda” category (17). Modern soda represents the only category innovation within the broader soda category since the introduction of diet or zero sugar sodas decades ago. Beverages within this novel category are often formulated with low-to no-added sugars, in addition to functional ingredients to support health benefits, such as soluble prebiotic fibers known to support gut or microbiome health.

Despite a healthful formulation of lower sugar and prebiotic fibers, there is no demonstrated effect on the postprandial glycemic impact of functional food and beverage products, suggesting a need for human trials that validate the glycemic response to these newly emerging “modern sodas (18)”. Thus, the primary aim of this human trial was to compare the acute effects of a lower-sugar, high-fiber prebiotic cola to a commercially available traditional cola on glucose excursions in response to the beverages alone and in combination with a carbohydrate-rich mixed lunch meal in free-living, generally healthy adults. To our knowledge, this is the first study to assess the postprandial impact of swapping traditional soda for modern prebiotic soda.

## METHODS

### Participants

Thirty men and women (18 – 65 y) with overweight or obesity (BMI: 25.0 – 34.9 kg/m^2^), but deemed otherwise healthy on the basis of medical history review with a study physician were eligible for study participation. Healthy participants with overweight or obesity were selected due to their elevated risk for cardiometabolic abnormalities, making them a relevant population for evaluating dietary interventions. Inclusion criteria included fasting glucose <126 mg/dL (confirmed by capillary fingerstick), controlled blood pressure (<140/<90 mm Hg), weight-stable (±4.5 kg within 90 d), and non-user of tobacco/nicotine (≥12 mo cessation) or marijuana (≥60 d cessation) products. Individuals were excluded if they had a diagnosis history of diabetes, eating disorders, gastrointestinal disorders, or any other clinically relevant conditions. Exclusionary medications and supplements included those known to affect blood glucose (e.g., diabetes-, mental health-, or thyroid-related medications), oral/injectable steroids (within 90 d), and daily fiber supplementation (within 30 d). Lifestyle exclusions included extreme dietary habits (e.g., ketogenic, vegan), current weight loss and/or muscle gain program, or shift workers with eating schedules that interfere with study design. Women currently pregnant (confirmed by pregnancy test at screening) or lactating were also excluded.

This study was carried out at Biofortis, Inc. (Addison, IL) in compliance with the protocol and in accordance with Good Clinical Practices (GCP), the applicable US Code of Federal Regulations (CFR), and the Declaration of Helsinki. The study protocol was approved by an Institutional Review Board (Sterling IRB, Atlanta, GA). Signed written informed consent was obtained from recruited individuals prior to participation. This trial was registered at ClinicalTrials.gov (NCT06427915).

### Experimental Design

In a free-living setting (i.e., at-home), participants consumed either a prebiotic soda (PREB) or a traditional soda (SODA) with or without a carbohydrate-rich meal as a daily lunch (PREB, SODA, PREB+CHO, SODA+CHO), in a randomized controlled crossover fashion for 4 consecutive test days. Test condition crossover sequence was counterbalanced following a Williams design with equal allocation to test sequence. The randomization schedule was prepared by a Biofortis statistician and uploaded to Medrio eCRF platform (Medrio, Inc. San Francisco, CA), with randomization number and associated sequence assigned upon eligibility confirmation. Participant screening included informed consent and eligibility assessment (e.g., medical history, inclusion/exclusion criteria, concomitant medication/supplements). Height, weight, vital signs (blood pressure and heart rate), and fasting capillary glucose by fingerstick, were measured under standard procedures. Upon successful screening, enrolled participants returned to clinic for a separate visit for continuous glucose monitor (CGM) application and an orientation on DexCom CGM and Cronometer diet record smart phone applications (*detailed in following sections*).

An overview of the experimental design is depicted in **Figure 1**. Each test day, participants were instructed to consume a free-living breakfast no later than 08:00. From 08:00 to time of experimental lunch (t = 0 min, 12:00 ± 30 min), participants were instructed to refrain from any food or beverages, with the exception of water. The experimental lunch included a 12 fl oz carbonated beverage as a modern prebiotic soda (PREB: OLIPOP^®^ Vintage Cola) or traditional soda (SODA: Original Coca-Cola^®^) (**Table 1**) with or without a plain bagel and cream cheese. This was an open-label pilot study and therefore not blinded. Participants were instructed to consume the lunch within 15 min. The next 3 h served as a post-meal observational period during which participants were instructed to refrain from any food or beverage (water allowed). Secondary outcomes of hunger and alertness were each assessed with visual analog scales (VAS) pre-meal (t = –30 min) and periodically during the post-meal period (t = 30, 120 and 180 min) with a ±10 min window. Text message reminders were sent for each VAS timepoint. After completion of the 180 min VAS, participants could resume independent habitual activities in accordance with intervention guidance.

**Figure 1.**
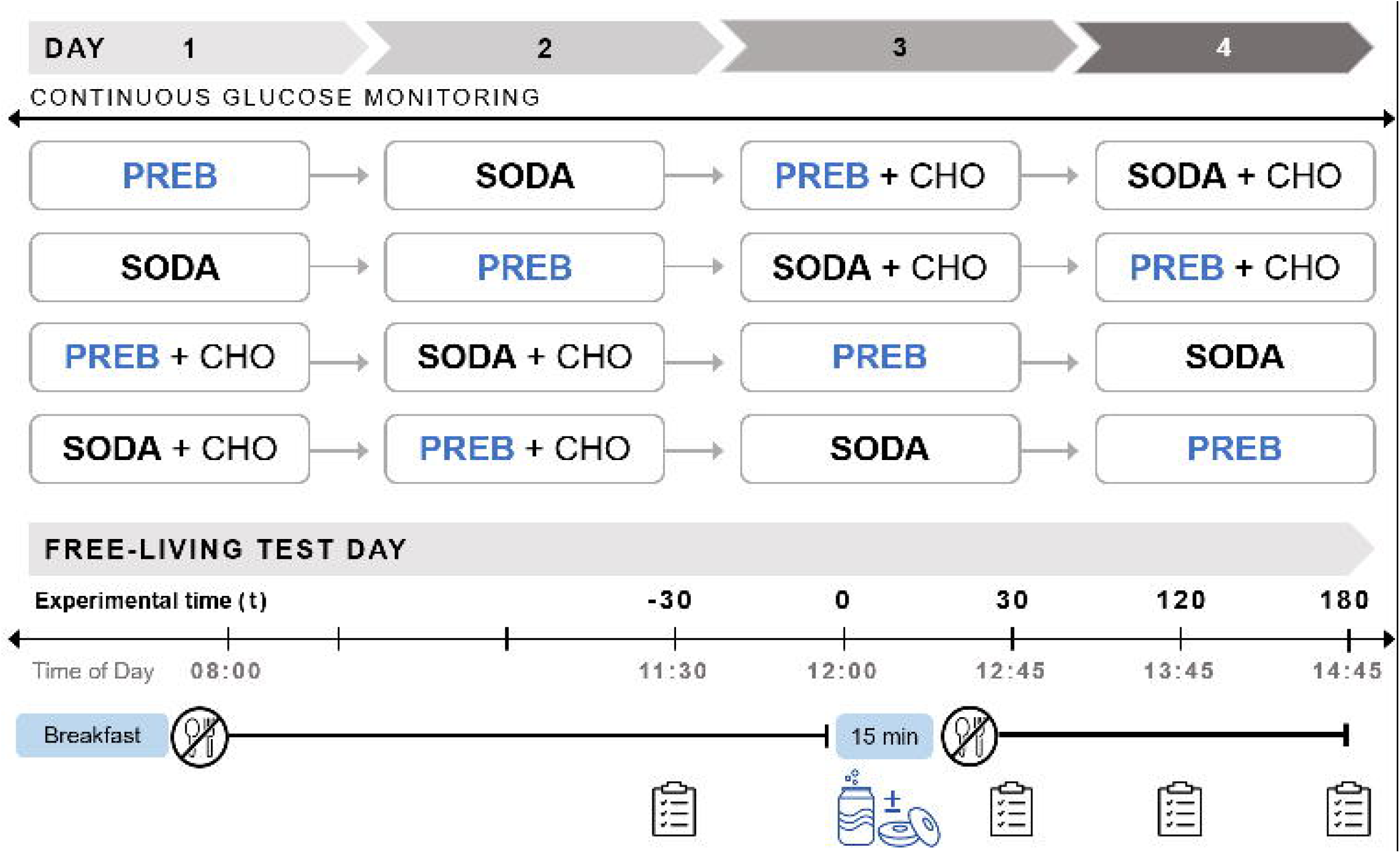
Experimental Design. Participants were randomized for a crossover 4-day intervention, counterbalanced for product sequence, consuming either a prebiotic soda (PREB) or traditional sugar-sweetened soda (SODA) ± carbohydrate-rich meal (CHO). Test days included a free-living breakfast (≤08:00), experimental product lunch (t=0, 12:00 – 12:15 ± 30 min), followed by a ∼ 3 h postprandial period. Appetite and alertness were assessed with a visual analog scale at t=-30, t=30, t=120, and t=180 (± 10 min). A continuous glucose monitor was active for the entirety of the intervention.

**Table 1.** Test lunch nutrient description.

| Nutrition | PREB | SODA | + CHO |
| --- | --- | --- | --- |
| Energy (kcal) | 30 | 140 | 354 |
| Fat (g) | 0 | 0 | 9.6 |
| Total Carbohydrate (g) | 11 | 39 | 54 |
| Dietary Fiber (g) | 6 | 0 | 2 |
| Total Sugars (g) | 3 | 39 | 7 |
| Protein (g) | 0 | 0 | 12 |

Nutrition information of test beverages (PREB: OLIPOP® Vintage Cola; SODA: Original Coca-Cola®) and the carbohydrate-rich meal (+ CHO) consisting of a plain bagel with cream cheese. The 4 test conditions included each beverage consumed with or without the meal (PREB, SODA, PREB+CHO, SODA+CHO) whereby nutrients consumed on “with meal” conditions equal the sum of the beverage and +CHO columns.

Volunteers were instructed to maintain their habitual diet and physical activity patterns throughout participation, but refrain from alcohol, vitamin C supplements, and fiber or prebiotic supplements. Vigorous physical activity was prohibited for the 24 h leading up to screening and each morning of test days throughout the remainder of the observation period (i.e., ≥3 h after lunch exercise was permissible).

### Continuous Glucose Monitoring

A DexCom G7 CGM (California, USA) was applied on the back of the arm above the elbow, in-clinic the day before testing. Participants downloaded the DexCom G7 smart phone app on their personal device and after a 30 min warm-up period, a working sensor was confirmed by glucose values updating within the app. Accordingly, participants were unblinded to their glucose values. Participants returned to the clinic after the 4-day test period for sensor removal and data capture.

The CGM measured blood glucose concentration approximately every 5 min. Data was imported into the Cronometer app as the primary source for all data analysis. Baseline blood glucose was defined as the mean glucose concentration across all test conditions from t ≥-30, ≤0 min. Postprandial timeframes included: 0-2 h postprandial (t >0, ≤120 min) and 2-3 h postprandial (t >120, ≤180 min). The primary outcome of postprandial 0-2 h incremental area under the curve (iAUC_0-120 min_) was calculated with the trapezoid method whereby t=0 was the average intra-individual baseline of each test condition, and t >0, ≤120 included all CGM-captured data points within the timeframe. Secondary glucose response outcomes included: maximum glucose concentration (C_max_), time to glucose C_max_ (T_max_), glucose rise (C_max_ relative to baseline), proportion of participants with sustained glucose dip (3 consecutive glucose values below baseline within the 2-3 h postprandial period), and proportion of participants with a 2-3 h mean glucose lower than baseline.

### Diet Records

Participants recorded all dietary intake with time of day, including test lunches, during the 4 test days on the Cronometer (Cronometer Software Inc, British Columbia, CAN) diet recording smart phone application. Secondary dietary outcomes included: second meal energy intake and timing (minutes elapsed from test lunch to next meal of ≥50 kcal) (19), as well as 24 h intake (daily totals from 4 h prior to breakfast to 20 h after).

### Statistical Analysis

Under the paired t-test assumption to measure potential differences in blood glucose iAUC, a sample of 26 subjects provides approximately 80% power to detect an effect size of 0.50 at the two-sided 0.05 significance level. A total of 30 subjects were randomized to allow for possible attrition, noncompliance, or missing data.

CGM related outcomes were evaluated with a mixed model containing fixed effect terms for test condition, test period, and test sequence with participant nested within test sequence as the random effect. In the event of model assumption violations (sensitivity analysis), the differences between test conditions were evaluated with the Wilcoxon sign rank test. Hunger and appetite ratings were evaluated with a mixed analysis of covariance (ANCOVA). The comparisons of interest included PREB and SODA with and without a meal (i.e., PREB vs SODA, and PREB+CHO vs SODA+CHO). No adjustment for multiple comparisons was planned, and therefore, each comparison was made at the 0.05 significance level maintaining the overall error rate at the 0.10. Sustained glucose dip (participant count and percentages) was compared between PREB and SODA with and without meal, with McNemar’s test. Statistical programming and analyses were performed using SAS^®^ (SAS Institute, Inc., North Carolina, USA), version 9.4, and R (R Core Team 2022) version 4.3.3. The *ncar* R package was used for calculation of the glucose iAUC, C_max_, and T_max_.

Review of the intent-to-treat (ITT) baseline glucose CGM data exhibited higher coefficient of variation (CV) than previous reports demonstrating a strong reproducibility and concordance between devices within participants of 3.7% median CV (IQR 1.7%–7.1%) (20). Baseline glucose CV across test days was >10% for 6 participants in our present cohort. The high variability may have been due to technical issues with a monitor, or unreported protocol deviations like medication use, physical exercise, and/or energy intake during the fasting window. All of these potential factors would make baseline data less reliable and thus, these 6 participants were excluded from CGM-derived outcomes in a sensitivity analysis. All other outcomes were analyzed with the ITT cohort as they were not CGM-derived measurements.

## RESULTS

### Participant characteristics

The flow of participants through the study is depicted in **Figure 2**. On average, our cohort (n=30) was middle-aged [mean ± standard deviation (SD) 46.5 ± 10.4 y] men and women with an overweight BMI (29.5 ± 2.6 kg/m^2^) (**Table 2**). The cohort was majority middle-aged white females. As discussed in the previous section, six participants were removed from postprandial (0-2 h) outcomes for a sensitivity analysis due to high variability in baseline blood glucose across test days. ITT baseline blood glucose was 96.8 ± 11.3 mg/dL [CV median (min, max) 5.7% (1.7%, 25.8%)]. Baseline blood glucose with participant removal after sensitivity analysis was 96.7 ± 9.9 mg/dL [CV 4.6% (1.7%, 9.5%)].

**Figure 2.**
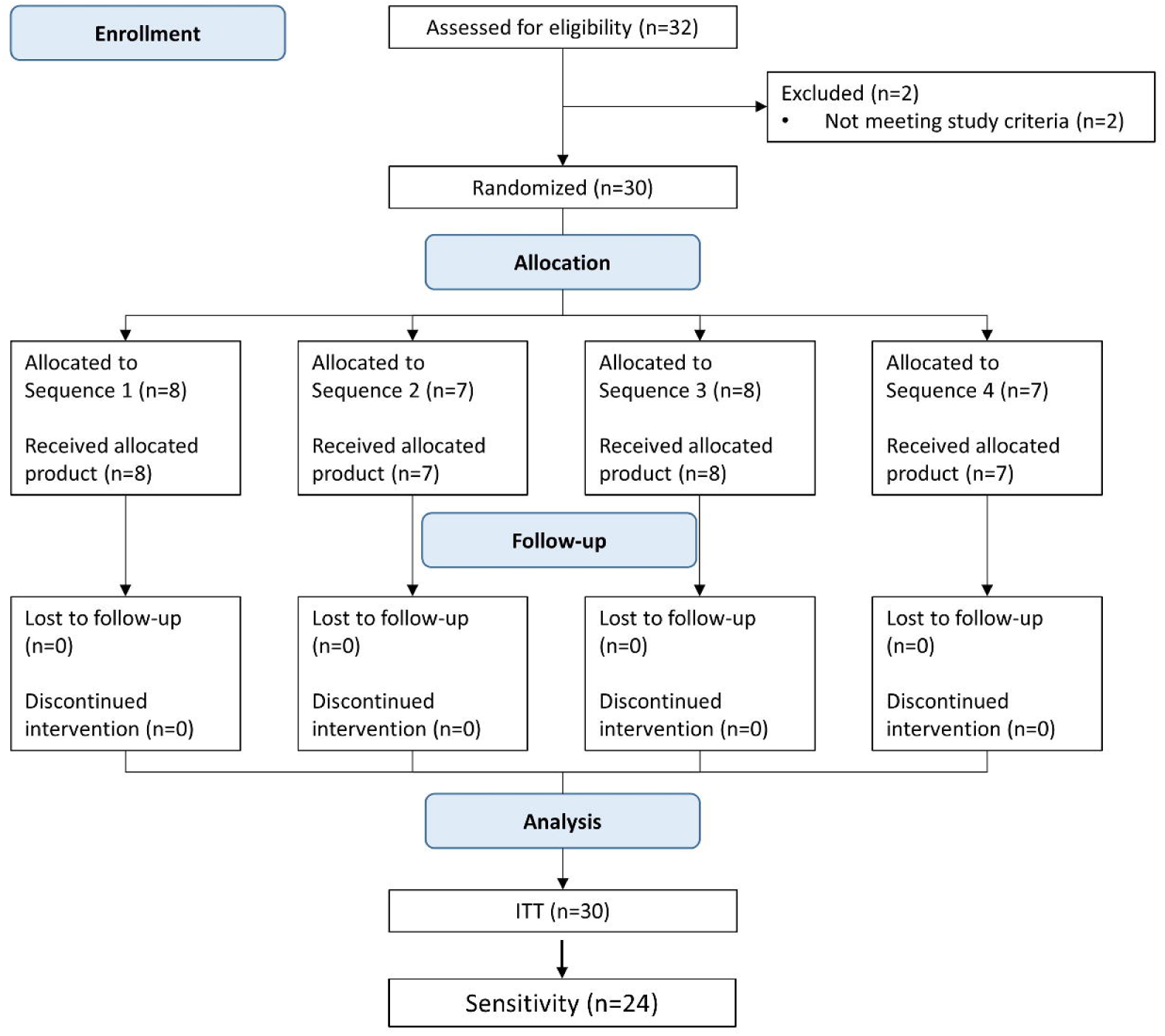
CONSORT Flow of Participants. CONSORT flow diagram illustrating number of individuals at each stage of the study.

**Table 2.**
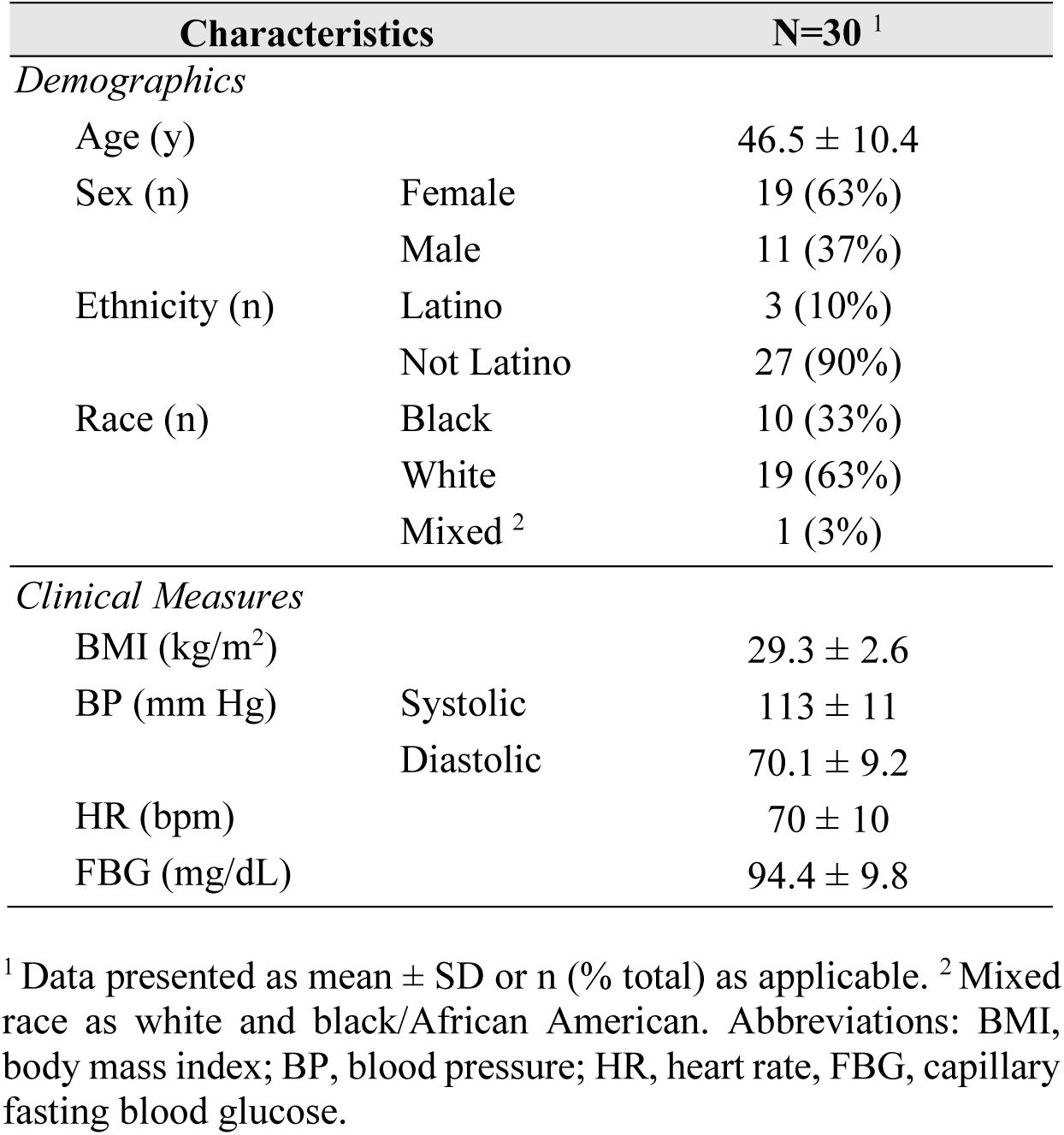
Participant characteristics.

### Postprandial (0-2 h) glucose is lower with prebiotic soda with or without a carbohydrate-rich meal

Postprandial (0-2 h) glucose response curves are illustrated in **Figure 3**. Glucose exposure (iAUC) was lower with PREB compared to SODA when consumed with a meal (p=0.032) and without a meal (p<0.001) (**Figure 4A**). In combination with a meal, C_max_ was also lower with PREB+CHO compared to SODA+CHO [MD: –9 mg/dL (95% CI: –27, 0), p=0.018]. Likewise, peak glucose concentration (C_max_) was lower with PREB alone compared to SODA alone [median difference (MD): –36 mg/dL (95% CI: –50, –22), p<0.001] (**Figure 4B**).

**Figure 3.**
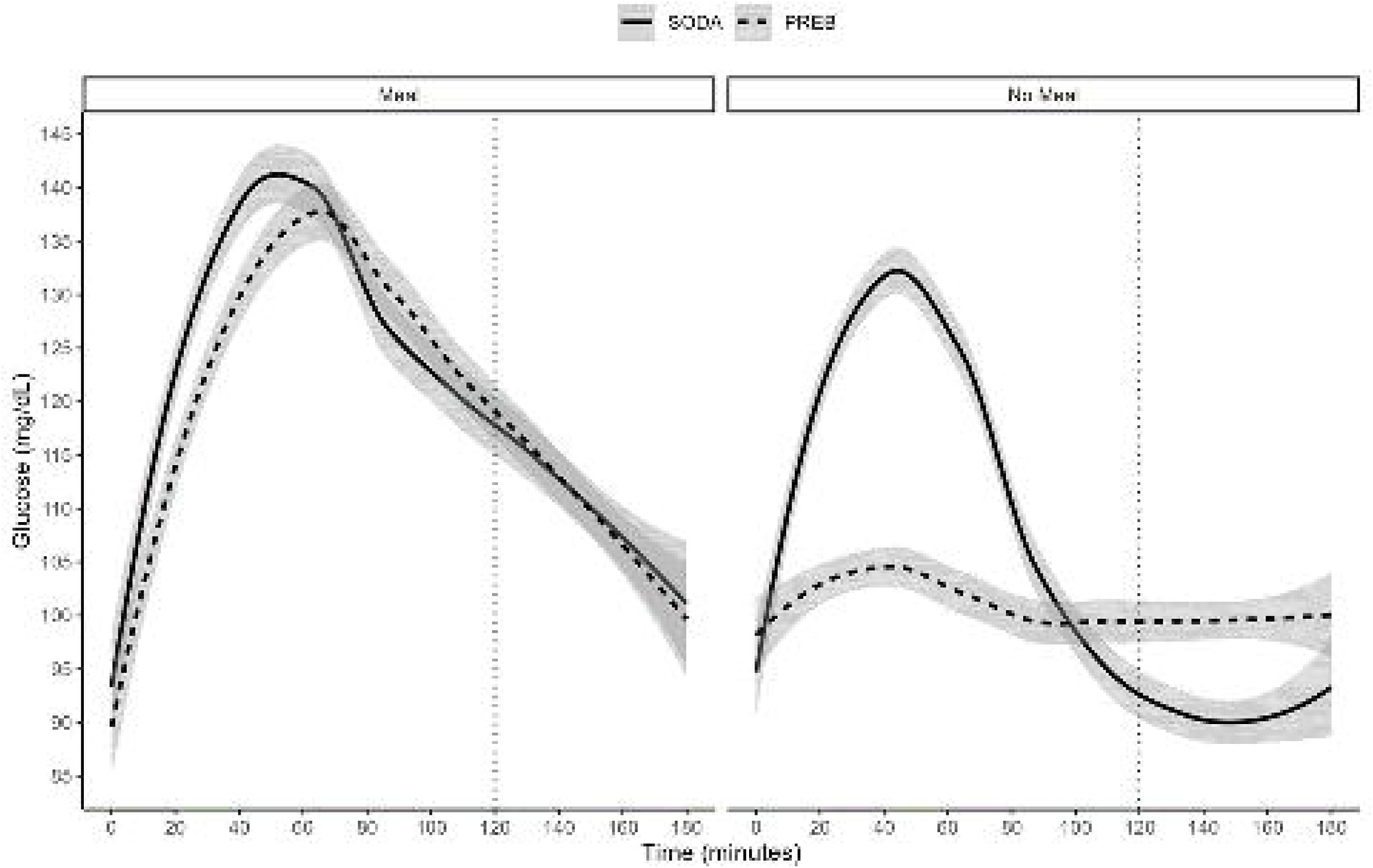
Postprandial glucose response to prebiotic or traditional soda with or without a meal. LOESS (locally estimated scatterplot smoothing) of blood glucose concentration with 95% confidence bounds, in response to a prebiotic soda (PREB) or traditional sugar-sweetened soda (SODA) separated by meal and no meal conditions. Time 0 is the average baseline glucose concentration over all test days, with the test lunch consumed immediately after. The dotted line represents 120 min post-test lunch, delineating the 0-2 h from 2-3 h postprandial periods.

**Figure 4.**
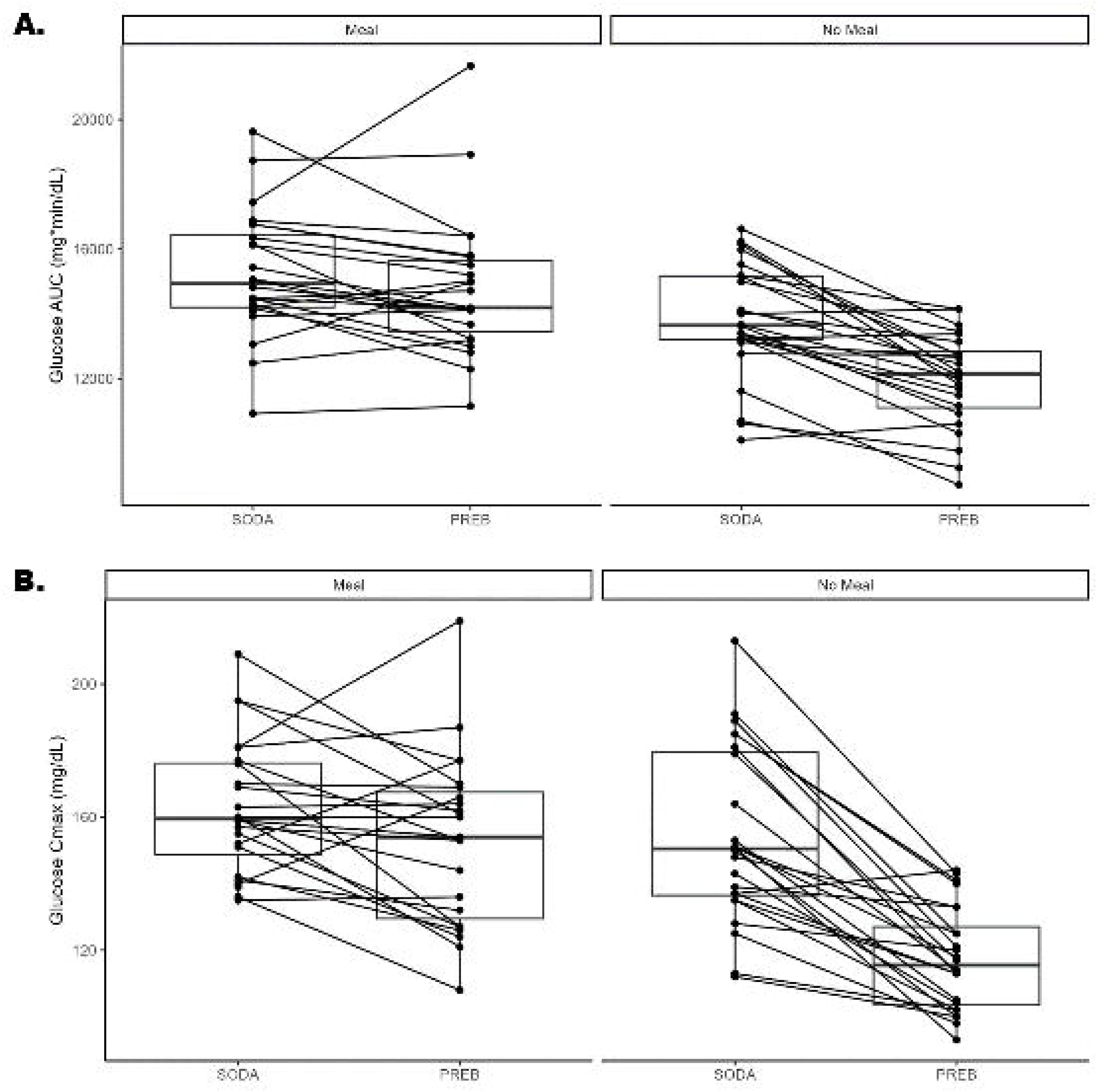
Postprandial (0-2 h) glucose AUC and C_max_ is lower with prebiotic soda. Postprandial (0-2 h) blood glucose **(A)** area under the curve (AUC) and **(B)** peak concentration (C_max_) in response to a prebiotic soda (PREB) or traditional sugar-sweetened soda (SODA) separated by meal and no meal conditions. Boxplots represent the distribution of responses under the given condition and the lines represent a single participant’s change between conditions.

Glucose rise was also lower in PREB than SODA both with a meal (MD: –10% (95% CI: –28, 0), p=0.025) or alone (MD: –38% (95% CI: –52, –23), p<0.001). Moreover, time to peak glucose levels (T_max_) were prolonged with PREB compared to SODA within a meal [MD: 13.5 min (95% CI: 5, 21), p=0.030]. Conversely, beverage choice did not alter T_max_ as a beverage alone (MD: –4.5 min (95% CI: –8, 5), p=0.25).

### Postprandial glucose dip (2-3 h) is unaffected by prebiotic soda

The proportion of participants with a sustained glucose dip was not different between PREB and SODA regardless of meal (meal: PREB: 17.2%, SODA: 20.7%, p=0.74; alone: PREB: p=0.13). Likewise, the proportion of participants who experienced a reduction in 2-3 h mean glucose compared to baseline was not significant between beverages (meal p=0.51; alone p=0.15).

### Prebiotic soda does not affect second meal timing nor energy intake

Total daily energy intake was not different between test conditions (main effect p=0.55). Timing and energy load of the next meal was not affected by PREB (**Table 3**). The main effect of second meal timing (≥50 kcal consumed) was not significant (p=0.17). On average, the second meal was consumed approximately 4 h after test conditions of beverage alone and approximately 4.5 to 5 h after test conditions of beverage with meal. The second meal energy intake on average ranged from approximately 300 to 430 kcal between all test conditions.

**Table 3.** Energy intake.

| Diet Records | Beverage alone |  | With meal |  | *P* |
| --- | --- | --- | --- | --- | --- |
|  | OLIPOP | SODA | OLIPOP | SODA |  |
| *Second Meal* |  |  |  |  |  |
| Time (h:min) | 3:54 ± 1:26 | 4:15 ± 2:04 | 5:08 ± 2:06 | 4:20 ± 1:29 | 0.17 |
| Energy (kcal) | 425 (212, 613) | 302 (156, 609) | 418 (256, 643) | 344 (170, 496) | 0.23 |
| *Daily Totals* |  |  |  |  |  |
| Energy (kcal/d) | 1330 (1030, 1590) | 1420 (1040, 1750) | 1210 (735, 1530) | 974 (736, 1470) | 0.55 |

Data rounded to 3 significant values and presented as mean ± SD or median (Q1, Q3) as applicable.

### Prebiotic soda does not affect perceived hunger nor alertness

The time course of patient-reported hunger and alertness VAS is illustrated in **Figure 5A-B**. Neither perceived hunger nor alertness was influenced by test condition. Pre-lunch (t=-30) hunger and alertness varied with a CV of 35.5% and 17.9%, respectively. Main effect of joint conditions [beverage (PREB × SODA) × meal (alone × CHO)] in the post meal (2-3 h) period was not significant for both hunger (p=0.55) and alertness (p=0.81).

**Figure 5.**
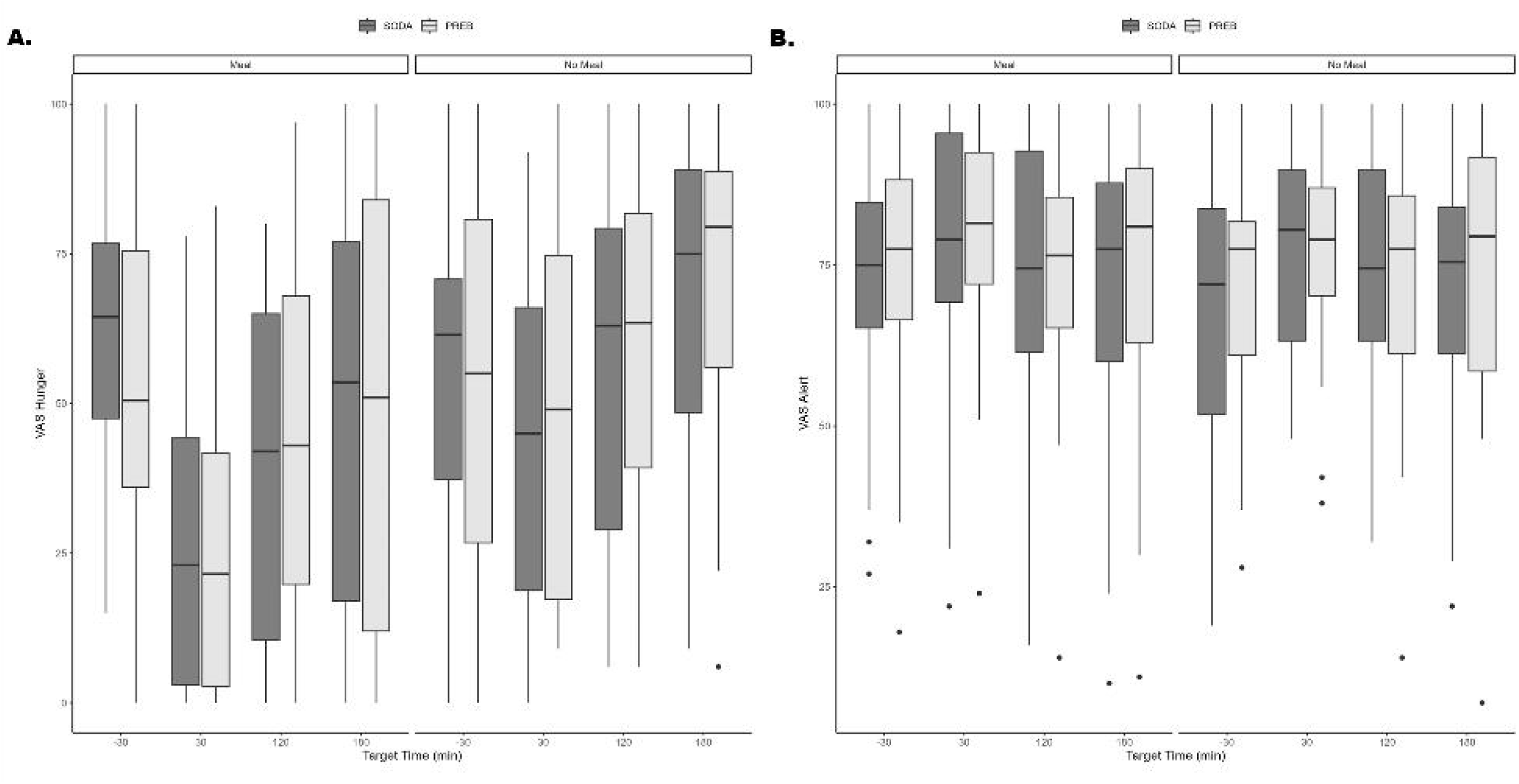
Prebiotic soda does not affect perceived hunger nor alertness. Time course of patient-reported **(A)** hunger and **(B)** alertness visual analog scales (VAS) in response in response to a prebiotic soda (PREB) or traditional sugar-sweetened soda (SODA) separated by meal and no meal conditions. Time –30 represents pre-lunch scores, with the test lunch consumed at t=0 min. Boxplots represent the distribution of response at each measured time point.

## DISCUSSION

This study assessed the impact of a prebiotic, lower sugar soda (PREB) on postprandial blood glucose compared to traditional sugar-sweetened soda. Generally healthy adults with overweight or class 1 obesity consumed prebiotic or traditional soda with and without a carbohydrate-rich meal. The prebiotic soda resulted in significantly lower postprandial glucose excursions (e.g., AUC, C_max_, and glucose rise over 0-2 h) compared to traditional soda when consumed alone or with a meal. Conversely, the late postprandial phase (i.e., 2-3 h) glucose dip was not different between beverages. Behavioral effects, such as second meal timing and energy intake, as well as perceived hunger or alertness, were not different with prebiotic soda compared to traditional. Overall, prebiotic soda, is a favorable beverage choice for managing postprandial glucose flux compared to traditional soda options.

The prebiotic soda evaluated in this study contains 28 (72%) fewer grams of total carbohydrate and 36 (77%) fewer grams of total sugars than the traditional soda (Table 1), categorizing PREB as a reduced sugar soda. This lower carbohydrate load is likely the primary contributing factor to lower glucose flux with PREB. In addition to its reduced sugar formulation, PREB contains 6g of dietary fiber (21% of the daily value), qualifying it for the FDA regulated “excellent source of fiber” nutrient content claim. Thus, it is worthwhile to note that comparable studies examining postprandial glucose dynamics observed favorable responses with prebiotic beverages independent of available CHO content (21, 22). Specifically, a recent study measured the 0-3 h postprandial glucose response to an inulin-dextrin tea supplement paired with a standardized mixed meal in adults with type 2 diabetes (22). The prebiotic blend reduced mixed-meal postprandial glucose and insulin AUC compared to the non-prebiotic tea beverage control. Moreover, routine prebiotic (resistant dextrin) beverage consumption (≥14 days) lowered blood glucose AUC and improved perceived satiety in the 0-2 h postprandial period compared to an iso-caloric and available CHO-matched beverage in generally healthy adults with normal-to-overweight BMI (21). Resistant dextrin is a component in the tested prebiotic soda bioactive blend, among other prebiotic fibers and plant botanicals. Thus, it is plausible to suspect that the prebiotic blend in PREB may serve as an additional modulator of blood glucose concentrations, in conjunction with the lower sugar content, compared to traditional soda.

Conversely, prolonged (>2 h) responses, like glucose dip, second meal effects, or perceived hunger or alertness, were not affected by the prebiotic soda. Second meal behavior (hunger, timing, and energy intake) is significantly associated with 2-3 h glucose dip (19), whereby greater dips in blood glucose predicts higher hunger, shorter time until next meal, and greater energy intake in the next meal. In present, a minor proportion of participants experienced a lower postprandial glucose dip than baseline regardless of condition, thus it is unsurprising that perceived hunger and second meal behavior was also not different across test meals. Translationally, these results highlight that despite the prebiotic soda being a lower calorie beverage alternative, consumption does not lead to earlier onset eating or overcompensation in energy intake with subsequent meals. Acute postprandial hunger was also unaffected in our current study. Prebiotic beverage supplementation has previously been shown to improve fasting and postprandial satiety, but significant differences were only observed after ≥14 days of twice daily prebiotic beverage consumption (21). However, BMI stratification identified that these satiety adaptations were exclusive to individuals with a normal BMI (21). The lack of effect of prebiotics on hunger in individuals with overweight or obese BMI, like in our cohort and others (21), may be a mere consequence of dose, whereby the beneficial effects of fiber intake may be relative to body weight. Alternatively, adiposity may affect the modulating capacity of the gut microbiome (23). Nevertheless, it is plausible to suspect that routinely choosing prebiotic soda over traditional variations may improve hunger.

Prebiotics have the potential to improve glycemic control through gut microbiome modulation, though this mechanism was not directly assessed in the present study. Intestinal microbe composition and corresponding byproducts [e.g., short-chain fatty acids, (SCFA)] are associated with diabetes risk in clinical observations (24) and have been shown to improve glycemic control in pre-clinical models (25). The bioactive blend of prebiotics and plant extracts incorporated in some modern soda formulations has been shown to promote *Bifidobacteria* growth and SCFA production in an *in vitro* gut fermentation model (26). Prebiotics not only increase abundance of beneficial microbes but have also been shown to reduce the abundance of potentially harmful microbes *in vivo* (27). For example, partially hydrolyzed guar gum supplementation, an ingredient in PREB, modulates microbiota populations, increasing those inversely associated with insulin resistance like *Verrucomicrobia*, and decreasing those positively associated with inflammation like *Dorea* and *Sutterella,* in healthy adults (28). Thus, our results provide a basis for future research opportunities to explore gut microbiome intermediaries in the role of cardiometabolic risk reduction with modern prebiotic sodas.

Various experimental parameters should be acknowledged when interpreting our results. Our study population was selected to be representative of the mean BMI observed within the U.S. adult population (29). Our free-living model in a generally healthy and representative BMI population enhances the translational efficacy of our results. Moreover, the test beverages are commercially available, ready-to-consume products, thus consumer implementation to swap traditional soda beverage of choice for a prebiotic soda is an accessible and feasible new health strategy to adopt. Likewise, our design of comparing sodas with and without a meal is also translational in that the benefits of prebiotics are achieved not only as a supplement (i.e., alone), but also when incorporated into an eating pattern (i.e., within a meal). Nevertheless, our study is not without limitations. Aligned with our free-living model of commercially available beverages, the products were open label. Lack of blinding poses a potential risk of bias for patient reported outcomes like VAS scores and diet records. As mentioned above, the differential carbohydrate load between test beverages is a likely contributing factor to the postprandial glycemic response. It is unclear how PREB may compare to reduced– or zero-sugar (without prebiotics) soda alternatives on postprandial glycemia. However, synthetic artificial sweeteners (e.g., aspartame as commonly incorporated in diet soda formulations) have been shown to modulate the gut microbiome (30), and are associated with type 2 diabetes risk (31). Thus ‘modern’ prebiotic sodas like PREB not only serve as lower sugar alternatives to SSBs, but also as naturally-sweetened (e.g., stevia), microbiome-friendly alternatives to diet beverages. Finally, the acute nature of this study limits conclusions about the long-term effects of prebiotic soda consumption on glycemic control or dietary behavior, warranting further research.

Overall, acute prebiotic soda consumption with and without a meal reduces postprandial glucose excursion, without affecting second meal behavior nor perceived hunger and alertness, compared to traditional soda. While it is not possible to isolate potential mechanisms, like reduced sugar content, higher fiber content, or gut microbiome modulation by prebiotic content, prebiotic soda nonetheless remains a favorable soda choice compared to traditional sugar-sweetened beverages.

## Data Availability

The data produced in the present study are not publicly shared

## ACKNOWLEDGEMENTS

This study was funded by the manufacturer of the prebiotic soda beverage, OLIPOP, Inc. CFM, TB, EM, VK, and CC were employees of Biofortis, Inc., a Contract Research Organization that received funding from OLIPOP, Inc. to conduct the study. NV, MJW, and CM were employees of OLIPOP, Inc., the study sponsor company. AMV was a consultant of OLIPOP, Inc. for research support at the sponsor’s request.

